# Prognostic performance of an AI-based recurrence risk model in clinically low-risk HR+/HER2- early breast cancer

**DOI:** 10.64898/2026.06.02.26354233

**Authors:** Cerise Tang, Dhruva Biswas, Chuwen Liu, Ken Zeng, Krzysztof J. Geras, Jan Witowski, Claudia Meurs, Pieter J. Westenend

**Author notes:** To Whom Correspondence Should be Addressed: Pieter J. Westenend.

## Abstract

**Objective:** Accurate prognostication of recurrence risk in early breast cancer is central for therapeutic decision-making, including identifying patients who may safely avoid adjuvant therapy. Here we evaluate an artificial intelligence (AI)-based method to improve risk stratification.

**Methods:** Ataraxis Breast CTX (ATX) is an AI test that integrates H&E-stained images with clinicopathologic features to predict risk of recurrence for individual patients. This study validates ATX in a dataset of 892 clinically low-risk patients from Dordrecht, Netherlands. Of the 892 patients, 299 did not receive adjuvant therapy. The discriminative performance of ATX was assessed using C-index and its stratification ability was evaluated by log-rank tests.

**Results:** ATX achieved a C-index of 0.71 and a 5-year AUC of 0.71, demonstrating strong discrimination. Among 299 patients who received no adjuvant therapy, ATX achieved a C-index and AUC of 0.78 and 0.81 respectively. ATX scores were used to stratify patients into risk groups. Notably, untreated and treated ATX low-risk patients had comparable 5-year recurrence-free survival (RFS) (untreated: RFS = 96%, 95% CI = 92-97%; treated: RFS = 96%, 95% CI = 93-97%) with identical 10-year RFS (86%, 95% CI = 83-92% for both), suggesting ATX low-risk status may identify a subgroup with favorable prognosis independent of treatment.

**Conclusion:** ATX provides robust prognostic stratification in an external cohort and identifies a subgroup of patients who did not receive systemic therapy with favorable observed outcomes. These results support prospective validation of ATX as a decision-support tool for adjuvant therapy de-escalation in HR+/HER2- early breast cancer.

## Introduction

The adjuvant treatment landscape for HR+/HER2- early breast cancer has evolved substantially in recent years, with the backbone or endocrine therapy now augmented by CDK4/6 inhibitors,^1,2^ PARP inhibitors for BRCA-mutated disease,^3^ and extended endocrine therapy regimens.^4,5^ In the current era, the standard of care for patients with early-stage HR+/HER2- breast cancer is surgery with or without radiotherapy and chemotherapy, followed by adjuvant endocrine therapy.^6^ Despite significant reductions in recurrence with adjuvant endocrine therapy, long-term follow-up beyond 20 years reveals recurrence rates of 27-37% for stage II disease and 46-57% for stage III disease.^7,8^

Alongside the expansion of standard of care regimens, genomic prognostic assays have gained prominence as decision-support tools, guiding adjuvant chemotherapy recommendations in the NCCN and ESMO clinical guidelines.^9,10^ Accurately estimating an individual’s risk of recurrence is essential to therapeutic decision-making. The consequences of overtreatment include substantial financial and physical burden with severe side effects that can dramatically reduce quality of life while undertreatment may lead to potentially preventable recurrence events. Thus, despite advances in genomic assays, improved prognostic risk stratification remains essential to optimize recurrence-free survival. Additionally, identification of patients at sufficiently low recurrence risk to safely forgo adjuvant therapy remains a critical and unmet clinical challenge, particularly in patients who have been deemed low-risk by conventional clinicopathologic criteria.

Artificial intelligence (AI) technologies applied to standard histopathology and clinical information could help address this unmet clinical need. If robustly validated, such an AI-based risk prediction platform could enable scalable, cost-effective, accurate prognostication.^11–18^ Ataraxis Breast CTX (ATX) is a multimodal AI test that integrates clinical information with morphological information extracted from H&E-stained whole-slide images to predict the 5-year risk of recurrence in early breast cancer patients.^19^ ATX leverages causal adjustment to learn to estimate recurrence risk under counterfactual therapy scenarios from observational data. Here, we evaluate ATX’s ability to predict recurrence in an external cohort of clinically low-risk early breast cancer patients.

## Methods

### Patient population and study design

A cohort of 892 patients diagnosed with early breast cancer at Albert Schweitzer Hospital in Dordrecht, the Netherlands, with available H&E slides and clinical information was assembled. All patients included in this study were diagnosed with HR+/HER2- early breast cancer. Slides were scanned using a Roche Ventana DP 200 scanner. The median follow-up time in this cohort was 9.1 years, as estimated by the reverse Kaplan-Meier method (**Figure S1)**. Treatment omission in this subgroup was consistent with Dutch national breast cancer guidelines, which do not recommend endocrine therapy for node-negative, low-risk tumors. For clarity, “untreated” patients throughout this manuscript denote patients who did not receive any adjuvant systemic therapy. Radiotherapy was not considered adjuvant systemic therapy.

### Development of Ataraxis Breast CTX

Ataraxis Breast CTX integrates pathology features, extracted from H&E-stained whole-slide images via a pathology foundation model,^20^ with clinical variables, to generate a patient-level representation. These representations are then used to estimate the probability of recurrence at 5 years under two counterfactual scenarios, when treated with endocrine therapy and chemoendocrine therapy. For this study, we use the endocrine therapy response prediction as a prognostic biomarker. ATX was trained on 9,141 patients from 12 institutions around the world. A pre-defined threshold of 0.065 (6.5%), reflecting the background 5-year recurrence risk in endocrine therapy-treated ER-positive early breast cancer,^21^ was used to dichotomize patients into high- and low-risk groups. No data from this dataset was used in developing ATX, and the model and threshold were locked prior to analyses in this dataset. Ataraxis Breast CTX is a class C2 predictive biomarker based on 2025 European Society of Medical Oncology (ESMO) biomarker classification criteria.^22^

### Survival Analysis

The primary endpoint in this study was recurrence-free survival (RFS), defined as duration from diagnosis to first recurrence or death and consistent with STEEP criteria.^23^ Across the full follow-up period, 128 recurrence events were observed (event rate = 14%). Overall survival was also assessed as a secondary endpoint. To measure accuracy of prognostic discrimination, we used two metrics: C-index^24–26^ and time-dependent AUC at 5 years as implemented in the scikit-survival package.^27^ Both metrics account for censored observations without requiring their exclusion, preserving the full analytic cohort. 95% confidence intervals were obtained using bootstrapping (1000 iterations).

Hazard ratios (HRs) were estimated to quantify the association between the score and recurrence risk. Continuous variables were standardized to two standard deviations to enable direct comparison of effect sizes with binary predictors, as proposed by Gelman.^28^ HRs were calculated using multivariate Cox models, adjusting for tumor stage, nodal stage, grade, age at diagnosis, receipt of adjuvant endocrine therapy, and receipt of adjuvant chemotherapy. Tumor stages 3 and 4 were collapsed into a single level to avoid sparse-level separation and ensure numerical stability. A second multivariate Cox model was fitted with only the untreated patients, adjusting for the same clinical variables.

Kaplan-Meier curves were estimated using the KaplanMeierFitter class from lifelines,^29^ confidence intervals were calculated using Greenwood’s formula, and p-values were calculated using log-rank tests. To compare treated and untreated ATX low-risk patients, HRs and 95% CI were estimated, with Kaplan-Meier curves generated as described above.

### Statistical Analysis

Distribution of clinical variables between treatment groups were tested using chi-square tests. Sample size calculations were not performed due to the retrospective nature of this cohort, all eligible patients were included. Statistical analyses were performed using Python 3.10 and R version 4.5.1. Data management and manipulation in Python were conducted using pandas (v1.5)^30^ and numpy.^31^

### Data visualization

All figures were made using matplotlib^32^ and ggplot2.^33^

## Results

### Cohort characteristics

Table 1 shows the clinicopathologic characteristics of the 892 patients in this study. The median age was 61 years old and 81% of patients were ≥ 50 years old. Tumors were predominantly stage T1 (71%), with only 2% classified as stage T3 or T4. The majority of patients were node-negative (71%) and presented with grade 2 tumors (56%). Histologically, 81% had invasive ductal carcinoma and 14% had invasive lobular carcinoma. Adjuvant therapy data were available for all patients. Most patients received radiotherapy (68%) and endocrine therapy (65%), while adjuvant chemotherapy was less common, administered to 30% of patients, consistent with the clinically low-risk nature of this cohort. The low rate of endocrine therapy was expected as Dutch guidelines support its omission in node-negative patients aged > 34 years with grade 1 tumors < 2 cm or grade 2–3 tumors < 1 cm, and in patients aged ≤ 35 years with grade 1 tumors < 1 cm.

**Table 1:** Characteristics of 892 clinically low-risk women with HR+/HER2- breast cancer. Variables are presented as n (% of patients).

| Characteristic | n (%) |
| --- | --- |
| **Age** |  |
| < 50 | 166 (18.6) |
| ≥ 50 | 726 (81.4) |
| **Tumor Stage** |  |
| T1 | 634 (71.1) |
| T2 | 241 (27.0) |
| T3 | 13 (1.5) |
| T4 | 4 (0.4) |
| **Nodal Stage** |  |
| N0 | 636 (71.3) |
| N1 | 249 (27.9) |
| N2 | 0 (0) |
| N3 | 0 (0) |
| NX | 7 (0.8) |
| **Grade** |  |
| 1 | 299 (33.5) |
| 2 | 499 (55.9) |
| 3 | 88 (9.9) |
| Unknown | 6 (0.7) |
| **ER Status** |  |
| Positive | 892 (100) |
| Negative | 0 (0) |
| **PR Status** |  |
| Positive | 791 (88.7) |
| Negative | 71 (8.0) |
| Unknown | 30 (3.3) |
| **HER2 Status** |  |
| Positive | 0 (0) |
| Negative | 892 (100) |
| **Subtype** |  |
| Lobular | 122 (13.7) |
| Ductal | 719 (80.6) |
| Mucinous | 22 (2.5) |
| Other | 29 (3.2) |
| **Endocrine Therapy** |  |
| Yes | 582 (65.2) |
| No | 310 (34.8) |
| **Chemotherapy** |  |
| Yes | 263 (29.5) |
| No | 629 (70.5) |
| **Radiotherapy** |  |
| Yes | 608 (68.2) |
| No | 284 (31.8) |
| **Surgery Type** |  |
| Lumpectomy | 595 (66.7) |
| Mastectomy | 297 (33.3) |

### ATX effectively stratifies clinically low-risk HR+/HER2- breast cancer

ATX demonstrated robust discriminative performance in predicting RFS, with a 5-year time-dependent AUC of 0.71 (95% CI = 0.64-0.77, **Figure S2a**) and a C-index of 0.71 (95% CI = 0.67-0.75). Next, to determine whether ATX is able to stratify patients by recurrence risk, we binarized patients into high- and low-risk groups. A total of 236 patients (26%) were classified as ATX high-risk while 656 (74%) were classified as low-risk. High-risk patients had a significantly lower 5-year RFS (87%, 95% CI = 82-91%) than low-risk patients (96%, 95% CI = 94-97%, **Figure 1a**). There was a statistically significant difference in RFS between the two groups across the follow-up period (log-rank p < 0.001). ATX high-risk patients had a markedly lower 10-year RFS of 64% (95% CI = 56-71%) compared to 88% (95% CI = 85-91%) in low-risk patients. ATX also demonstrated strong discriminative performance in predicting OS, with a 5-year time-dependent AUC of 0.73 and C-index of 0.72 (**Figure S3a**). The difference in OS between high- and low-risk groups was statistically significant (p < 0.001, **Figure S3c**). These results demonstrate that ATX effectively stratifies patients by recurrence risk beyond the 5-year window.

**Figure 1:**
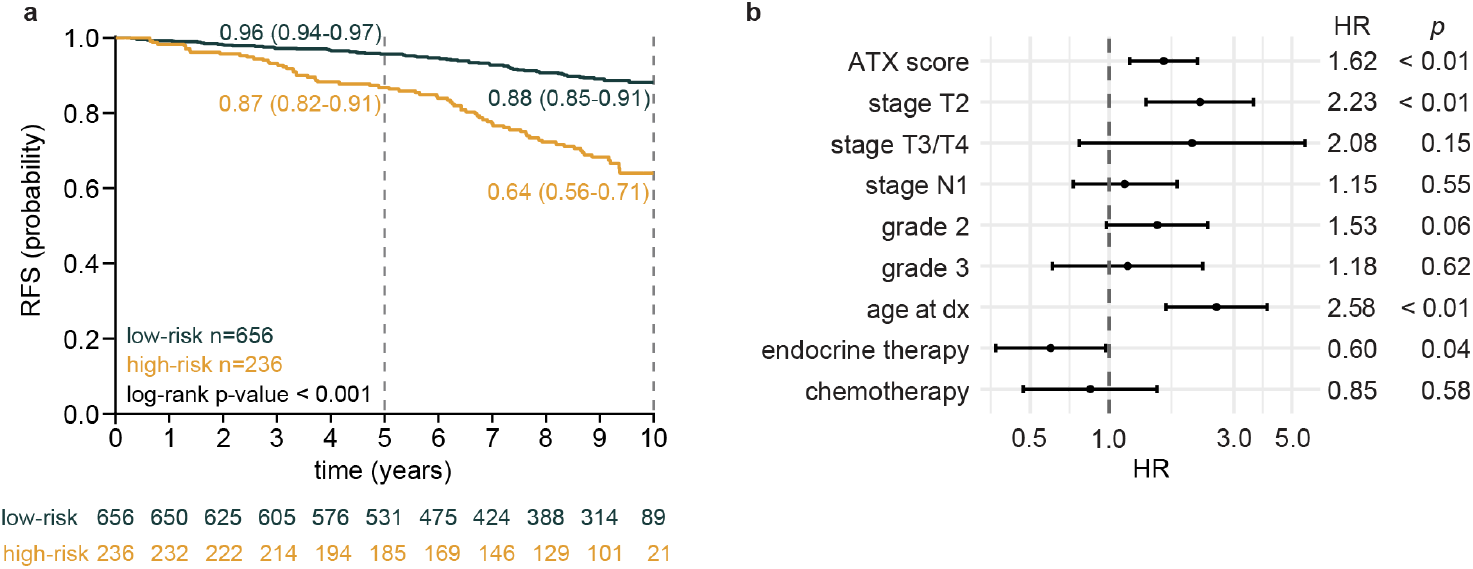
External validation of ATX prognostic performance in a clinically low-risk cohort. a) Kaplan-Meier curves illustrating recurrence-free survival stratified by ATX risk category, with 5 and 10-year estimates and corresponding 95% confidence intervals. Statistical significance of differences between ATX risk categories was determined using a log-rank test. Numbers below represent patients at risk at each time point. b) Multivariate Cox regression model demonstrating that ATX score (HR per 1 SD change in score) is associated with recurrence-free survival independent of clinical variables and adjuvant therapy received.

### ATX remains prognostic after adjusting for clinical variables

ATX captures signals beyond clinical variables using pathology features derived from H&E-stained whole-slide images extracted using a pathology foundation model.^20^ To demonstrate this, we fitted a multivariate Cox model adjusted for tumor stage, nodal stage, grade, age at diagnosis, receipt of adjuvant endocrine therapy, and receipt of adjuvant chemotherapy. ATX score remained independently associated with RFS (HR = 1.62, 95% CI = 1.20-2.18, Wald test p < 0.001; **Figure 1b**). In addition, stage T2, age at diagnosis, and receipt of adjuvant endocrine therapy were also independently associated with RFS in the multivariate model (p < 0.05). ATX was also independently associated with OS (HR = 1.52, 95% CI = 1.08-2.13, Wald test p = 0.02, **Figure S3d**).

### ATX stratifies prognosis for clinically low-risk patients who did not receive adjuvant therapy

In the 892 patient cohort, 299 patients did not receive adjuvant systemic endocrine therapy, chemotherapy, or targeted therapy. As expected, these patients had more favorable clinicopathologic features than treated patients. Specifically, patients without systemic therapy were predominantly T1 (93.0% vs 60.0%, p < 0.001, **Figure 2a**), node-negative (94.9% vs 59.4%, p < 0.001, **Figure 2b**), and low grade (64.5% vs 18.5%, p < 0.001, **Figure 2c**). Consequently untreated patients were also more likely to be classified as ATX low-risk (90.3% vs 65.1%, p < 0.001, **Figure 2d**). The full score distributions for treated and untreated samples are presented in **Figure S4**.

**Figure 2:**
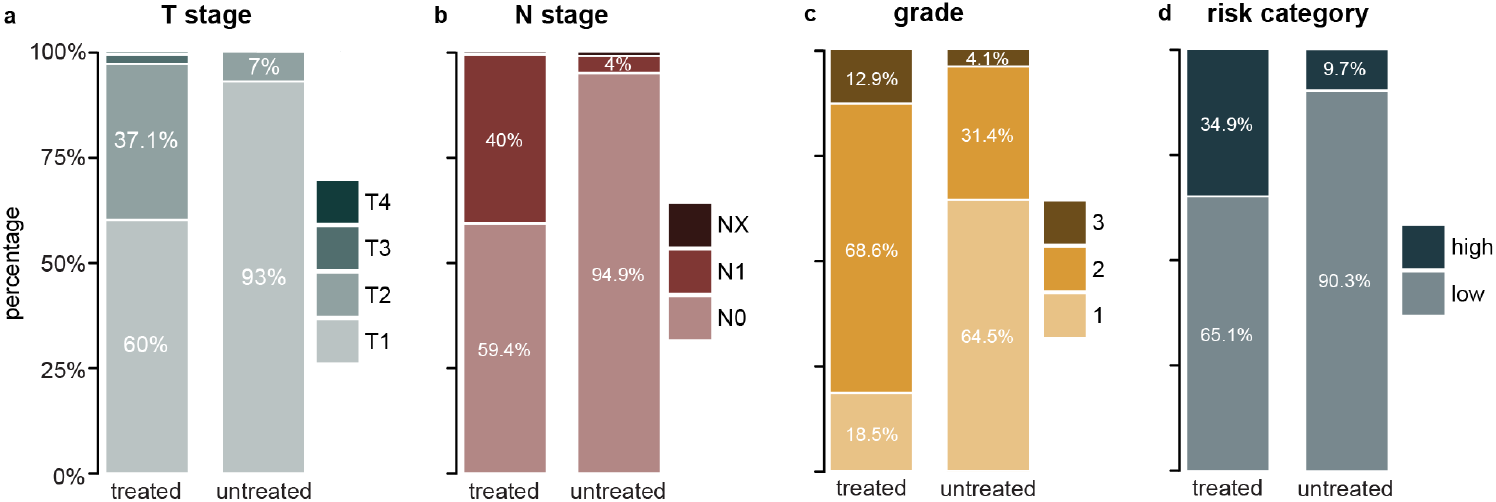
Clinicopathological differences between treated and untreated patients reflect treatment selection patterns. Stacked barplots comparing the distribution of patients with different a) tumor stages, b) nodal stages, c) grade, and d) ATX risk category between treated and untreated patients.

To determine whether ATX meaningfully stratifies recurrence risk within untreated patients, we next evaluated its prognostic performance in this subgroup. ATX demonstrated a 5-year time-dependent AUC of 0.81 (95% CI = 0.70-0.90), indicating strong discriminative performance for RFS (**Figure S2b**). The C-index in this cohort was 0.78 (95% CI = 0.70-0.84). When stratified into risk categories by ATX, 29 patients (10%) were classified as high-risk and 270 patients (90%) were classified as low-risk. Of the low-risk patients, 28 (10%) experienced an event compared to 15 (52%) of high-risk patients. ATX high-risk patients had a significantly lower 5-year RFS (64%, 95% CI = 44-79%) than low-risk patients (96%, 95% CI = 92-97%, **Figure 3a**). The two groups are statistically significantly different in RFS across the follow-up period (log-rank p < 0.001). At 10 years, ATX high-risk patients without adjuvant therapy had a significantly lower RFS of 46% (95% CI = 27-63%) compared to 88% (95% CI = 83-92%) in untreated low-risk patients. Additionally, ATX demonstrated discriminative ability for OS with a 5-year time-dependent AUC of 0.86 and C-index of 0.81 (**Figure S3b**). ATX also significantly stratified OS of high- and low-risk patients (log-rank p < 0.001, **Figure S3e**). However, the estimates for high-risk patients should be interpreted with caution given the small number of patients classified as high-risk, as demonstrated by the wide confidence intervals.

**Figure 3:**
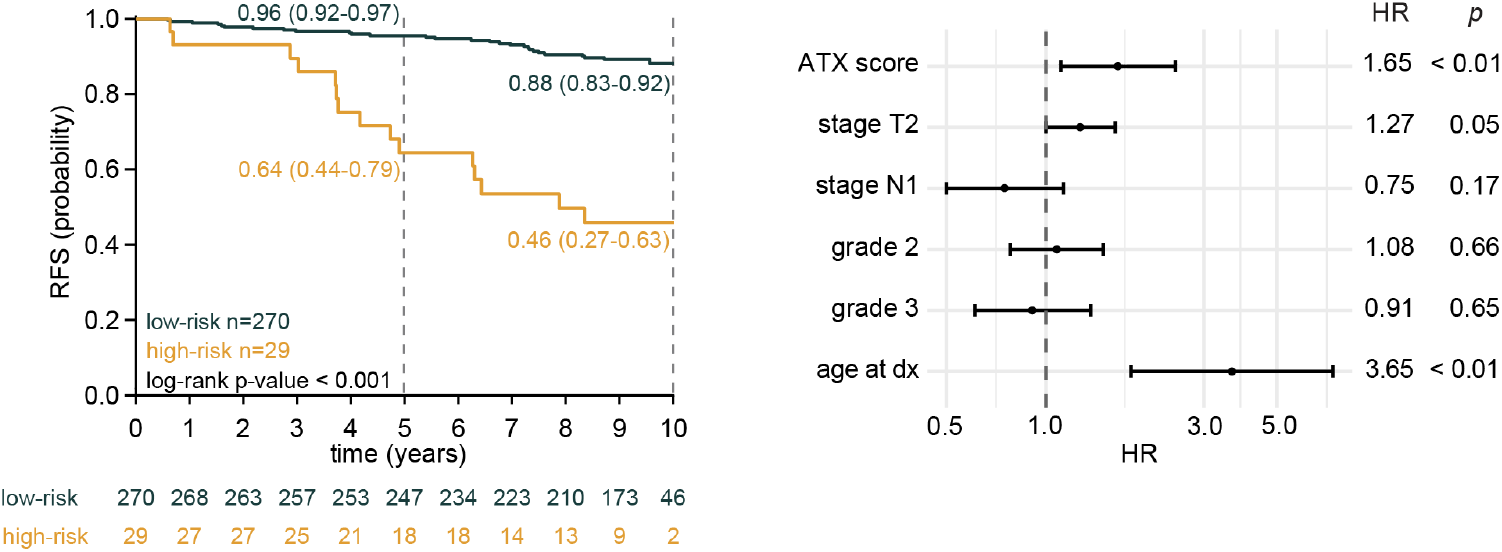
ATX effectively stratifies a cohort of untreated HR+/HER2- patients. a) Kaplan-Meier plot of recurrence-free survival with 5 and 10-year estimates and corresponding 95% confidence intervals shown. b) Multivariate Cox regression model demonstrates ATX score remains significantly associated with recurrence-free survival independent of clinical variables.

Finally, we assessed whether ATX was independently associated with RFS after adjusting for clinical variables. In a multivariable Cox model including tumor stage, nodal stage, grade, and age at diagnosis, ATX score remained independently associated with RFS (HR = 1.65, 95% CI = 1.11-2.46, Wald test p = 0.01; **Figure 3b**). ATX was also independently associated with OS (HR = 1.61, 95% CI = 1.05-2.47, Wald test p = 0.03, **Figure S3f**).

### ATX identifies untreated patients with outcomes comparable to treated patients

To evaluate whether ATX identifies untreated patients with outcomes not worse than treated patients, we compared recurrence-free outcomes between treated and untreated patients within the ATX-stratified low-risk group. Recurrence-free outcomes were similar between treated and untreated ATX low-risk patients. Five-year RFS in treated patients (n = 386) was 96% (95% CI = 93-97%) and 96% (95% CI = 92-97%) in untreated low-risk patients (n = 270). This pattern was consistent at 10 years where ATX low-risk patients who received no adjuvant therapy achieved an RFS identical to that of treated low-risk patients (10-year RFS = 88%, 95% CI = 83-92%), though low at-risk numbers and event counts lead to wider confidence intervals in untreated patients. Kaplan-Meier curves between treated and untreated patients had overlapping confidence intervals (log-rank test p = 0.94, **Figure 4**). Similarly, the log-rank test did not detect a significant difference in OS between treated and untreated patients (log-rank p = 0.36, **Figure S3g**), though this does not imply equivalence. A Cox proportional hazards model yielded an observed HR of 1.08 (95% CI = 0.64-1.80, p = 0.78) for untreated versus treated patients. Given the observational and exploratory nature of this analysis, combined with the limited number of events (28 and 30 in untreated and treated groups respectively), the comparison was underpowered as the hazard ratio estimate (HR = 1.08, 95% CI: 0.64–1.80) was compatible with a wide range of effect sizes, from a meaningful benefit to substantial harm of treatment omission. Formal non-inferiority testing was therefore not feasible and would require a prospectively designed trial with pre-specified margins and adequate event accrual.

**Figure 4:**
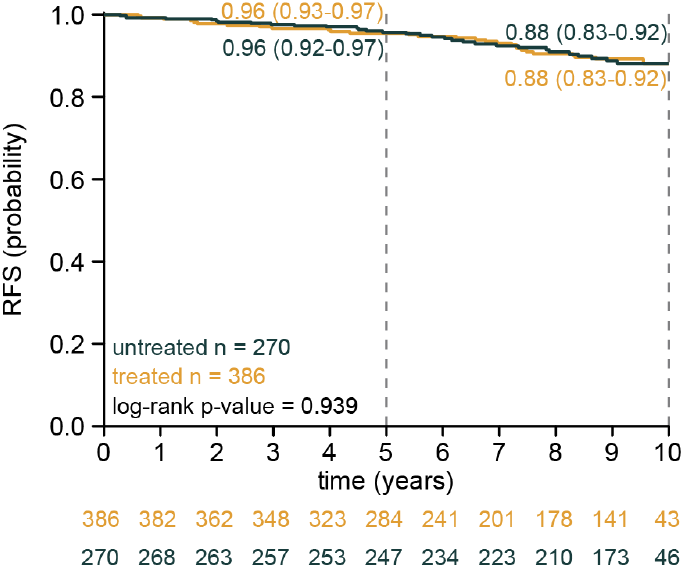
ATX low-risk patients have similar survival regardless of adjuvant therapy receipt. Kaplan-Meier plot of recurrence-free survival in ATX-stratified low-risk patients (n = 656). Estimates of recurrence-free survival at 5 and 10 years with their respective 95% confidence intervals are shown. The log-rank test did not detect a statistically significant difference between treatment groups (p = 0.94).

## Discussion

In this study, we show that ATX, a multimodal AI test that utilizes H&E images and clinical variables, identifies a subgroup of clinically low-risk HR+/HER2- breast cancer patients with excellent outcomes despite omission of adjuvant systemic therapy. Specifically, ATX-stratified low-risk patients achieved a 5-year RFS of 96% when untreated, comparable to that observed in treated low-risk patients (96%). While untreated patients had more favorable clinicopathologic features, ATX further stratified this already low risk group. We observed no absolute difference in 10-year RFS between treated and untreated ATX low-risk patients, with overlapping confidence intervals, though this analysis was not powered to establish non-inferiority. Notably, this low event count itself reflects the favorable biology ATX identifies, and equally precludes formal conclusions of equivalence between treated and untreated patients.

The results presented here are particularly relevant in the context of ongoing efforts to optimize treatment selection in early breast cancer, with increasing emphasis on minimizing overtreatment. While clinicopathologic criteria are currently used to identify patients at relatively low-risk, our results suggest ATX may further refine this group, isolating a subset with low observed recurrence rates. Importantly, while these findings do not establish that adjuvant therapy may be safely omitted, they support the hypothesis that a subset of patients may derive minimal benefit from treatment.

Beyond this clinically relevant subgroup, ATX demonstrated consistent prognostic performance in the full cohort with a C-index and time-dependent AUC of 0.71. By leveraging a pathology foundation model, ATX extracts morphological features from routinely-collected H&E images, presenting a scalable and cost-effective alternative to existing prognostic tools, such as genomic assays. Further, unlike other AI tests,^17^ ATX demonstrated prognostic utility across all histological subtypes, broadening its potential clinical applicability.^18^ The independent prognostic utility of ATX beyond conventional clinical variables was supported by its hazard ratios in both the full cohort (HR = 1.62, Wald test p < 0.001) and the untreated cohort (HR = 1.65, Wald test p = 0.01), demonstrating its utility as an independent prognostic indicator. Importantly, ATX remained independently significant after adjusting for tumor stage, demonstrating that it captures prognostic information beyond what staging alone provides.

In the full HR+/HER2- Dordrecht cohort, T2 stage yielded a hazard ratio of 2.23 relative to T1, substantially higher than the risk ratio of approximately 1.48 derived from the weighted 20-year recurrence rates reported by Pan et al.^7^ Age showed a similarly large effect (HR = 2.58), consistent with the cohort’s wide age distribution (IQR: 51-78) and the substantial differences in baseline risk and competing mortality. Although conventional prognostic factors carry larger-than-expected effect sizes in this cohort, ATX retained independent prognostic value.

When interpreting the results of this study, limitations that should be considered include the retrospective nature of this analysis. This introduces the potential for selection bias, particularly in the untreated subgroup where the decision to withhold adjuvant therapy was influenced by favorable clinicopathologic features. Confounding of this nature was evidenced by the significant differences in stage, grade, and nodal status between treated and untreated patients, however we note that the study’s main results were robust in multivariate analyses accounting for these features. Second, the small sample size of high-risk untreated patients (n=29) yielded wide confidence intervals in the estimation of long-term outcomes, motivating the need for validation in larger cohorts.

In conclusion, this external validation study demonstrates that ATX functions as a prognostic instrument in clinically low-risk HR+/HER2- early breast cancer. The comparable long-term outcomes observed between ATX low-risk untreated patients and ATX low-risk treated patients support the potential of ATX as a scalable, AI-enabled decision-support tool for the de-escalation of adjuvant therapy.

## CRediT authorship contribution statement

**Cerise Tang:** conceptualization, methodology, software, validation, formal analysis, investigation, data curation, writing - original draft, writing - review and editing, visualization, project administration. **Dhruva Biswas:** conceptualization, writing - original draft, writing - review and editing. **Chuwen Liu:** methodology, writing - review and editing. **Ken Zeng:** methodology, software, data curation, writing - review and editing. **Krzysztof J. Geras:** conceptualization, resources, methodology, writing - review and editing, project administration, supervision. **Jan Witowski:** conceptualization, resources, writing - review and editing, project administration, supervision. **Claudia Meurs:** conceptualization, writing - review and editing, supervision. **Pieter J. Westenend:** conceptualization, resources, writing - review and editing, project administration, supervision.

## Data Availability

The dataset analyzed in this study is private or proprietary and is not publicly available.

## Code Availability

Ataraxis Breast CTX is available for non-commercial research use upon reasonable request. The test is also commercially available for clinical use through Ataraxis AI. A Jupyter notebook to reproduce the analyses presented in this study is available upon request.

## Declaration of Competing Interests Statement

CT, DB, CL, KZ, KJG, and JW are equity holders of Ataraxis AI. All other authors declare no competing interests.

## Statement of Ethics

This study was performed under a data use agreement between Stichting Albert Schweitzer Ziekenhuis and Ataraxis AI, and in accordance with the General Data Protection Regulation (GDPR) and other applicable local data protection legislation. The institutional review board of Stichting Albert Schweitzer Ziekenhuis approved the use of human patient specimens and associated clinical data for this study, with a waiver of informed consent.

## Funding Sources

This study was funded by Ataraxis AI.

## References

1. Crown, J. et al. Adjuvant ribociclib plus nonsteroidal aromatase inhibitor therapy in patients with HR-positive/HER2-negative early breast cancer: 5-year follow-up of NATALEE efficacy outcomes and updated overall survival. ESMO Open 10, 105858 (2025).

2. Johnston, S. R. D. et al. Abemaciclib plus endocrine therapy for hormone receptor-positive, HER2-negative, node-positive, high-risk early breast cancer (monarchE): results from a preplanned interim analysis of a randomised, open-label, phase 3 trial. Lancet Oncol. 24, 77–90 (2023).

3. Tutt, A. N. J. et al. Adjuvant olaparib for patients with BRCA1-or BRCA2-mutated breast cancer. N. Engl. J. Med. 384, 2394–2405 (2021).

4. Gray, R. G. et al. aTTom: Long-term effects of continuing adjuvant tamoxifen to 10 years versus stopping at 5 years in 6,953 women with early breast cancer. J. Clin. Oncol. 31, 5–5 (2013).

5. Davies, C. et al. Long-term effects of continuing adjuvant tamoxifen to 10 years versus stopping at 5 years after diagnosis of oestrogen receptor-positive breast cancer: ATLAS, a randomised trial. Lancet 381, 805–816 (2013).

6. Pistilli, B., Lohrisch, C., Sheade, J. & Fleming, G. F. Personalizing adjuvant endocrine therapy for early-stage hormone receptor-positive breast cancer. Am. Soc. Clin. Oncol. Educ. Book 42, 1–13 (2022).

7. Pan, H. et al. 20-year risks of breast-cancer recurrence after stopping endocrine therapy at 5 years. N. Engl. J. Med. 377, 1836–1846 (2017).

8. Slamon, D. et al. Ribociclib plus endocrine therapy in early breast cancer. N. Engl. J. Med. 390, 1080–1091 (2024).

9. Loibl, S. et al. Early breast cancer: ESMO Clinical Practice Guideline for diagnosis, treatment and follow-up. Ann. Oncol. 35, 159–182 (2024).

10. Gradishar, W. J. et al. Breast Cancer, version 3.2024, NCCN Clinical Practice Guidelines in oncology. J. Natl. Compr. Canc. Netw. 22, 331–357 (2024).

11. Bidard, F.-C. et al. Identifying patients with low relapse rate despite high-risk estrogen receptor-positive/human epidermal growth factor receptor 2-negative early breast cancer: Development and validation of a clinicopathologic assay. J. Clin. Oncol. 43, 3090–3101 (2025).

12. Boehm, K. M. et al. Multimodal histopathologic models stratify hormone receptor-positive early breast cancer. Nat. Commun. 16, 2106 (2025).

13. Su, Z. et al. BCR-Net: A deep learning framework to predict breast cancer recurrence from histopathology images. PLoS One 18, e0283562 (2023).

14. El Agouri, H. et al. Assessment of deep learning algorithms to predict histopathological diagnosis of breast cancer: first Moroccan prospective study on a private dataset. BMC Res. Notes 15, 66 (2022).

15. Chen, Y. et al. Computational pathology improves risk stratification of a multi-gene assay for early stage ER+ breast cancer. NPJ Breast Cancer 9, 40 (2023).

16. Shamai, G. et al. Deep learning on histopathological images to predict breast cancer recurrence risk and chemotherapy benefit: a multicentre, model development and validation study. Lancet Oncol. 0, (2026).

17. Westenend, P. J. et al. External validation of precisebreast, a digital prognostic test for predicting breast cancer recurrence, in an early-stage cohort from the Netherlands. Breast Cancer Res. 27, 152 (2025).

18. Witowski, J. et al. Multi-modal AI for comprehensive breast cancer prognostication. Nat. Commun. 1–16 (2026).

19. Estimating individual benefit from adjuvant chemotherapy in hormone receptor–positive breast cancer using causal multimodal artificial intelligence. - ASCO. https://www.asco.org/abstracts-presentations/265844.

20. Cappadona, J. et al. Squeezing performance from pathology foundation models with chained hyperparameter searches. in NeurIPS 2024 Workshop: Self-Supervised Learning - Theory and Practice (2024).

21. Early Breast Cancer Trialists’ Collaborative Group. Electronic address. Reductions in recurrence in women with early breast cancer entering clinical trials between 1990 and 2009: a pooled analysis of 155 746 women in 151 trials. Lancet 404, 1407–1418 (2024).

22. Aldea, M. et al. ESMO basic requirements for AI-based biomarkers in oncology (EBAI). Ann. Oncol. 0, (2025).

23. Tolaney, S. M. et al. Updated Standardized Definitions for Efficacy End Points (STEEP) in adjuvant breast cancer clinical trials: STEEP version 2.0. J. Clin. Oncol. 39, 2720–2731 (2021).

24. Harrell, F. E., Jr, Lee, K. L. & Mark, D. B. Multivariable prognostic models: issues in developing models, evaluating assumptions and adequacy, and measuring and reducing errors. Stat. Med. 15, 361–387 (1996).

25. Harrell, F. E., Jr, Lee, K. L., Califf, R. M., Pryor, D. B. & Rosati, R. A. Regression modelling strategies for improved prognostic prediction. Stat. Med. 3, 143–152 (1984).

26. Harrell, F. E., Jr, Califf, R. M., Pryor, D. B., Lee, K. L. & Rosati, R. A. Evaluating the yield of medical tests. JAMA 247, 2543–2546 (1982).

27. Pölsterl, S. scikit-survival: A Library for Time-to-Event Analysis Built on Top of scikit-learn. Journal of Machine Learning Research 21, 1–6 (2020).

28. Gelman, A. Scaling regression inputs by dividing by two standard deviations. Stat. Med. 27, 2865–2873 (2008).

29. Davidson-Pilon, C. lifelines: survival analysis in Python. Journal of Open Source Software 4, 1317 (2019).

30. Team, T. P. D. Pandas-Dev/pandas: Pandas. (Zenodo, 2020). doi:10.5281/zenodo.3509134.

31. Harris, C. R. et al. Array programming with NumPy. Nature 585, 357–362 (2020).

32. Hunter, J. D. Matplotlib: A 2D Graphics Environment. Comput. Sci. Eng. 9, 90–95 (2007).

33. Wickham, H. ggplot2: Elegant Graphics for Data Analysis. Preprint at https://ggplot2.tidyverse.org (2016).

