## Supplemental Figures for "Prognostic performance of an AI-based recurrence risk model in clinically low-risk HR+/HER2- early breast cancer"

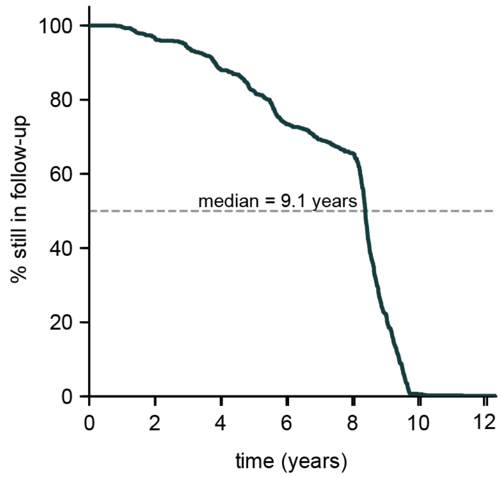


**Figure S1: Censoring distribution of the 892 patient cohort over time.** The y-axis represents the estimated percentage of patients who remain under active follow-up, and the x-axis represents time in years. The curve was generated using a reverse Kaplan-Meier estimator, where censoring is treated as the event of interest. The dotted gray line represents the time at which 50% of patients have been censored.

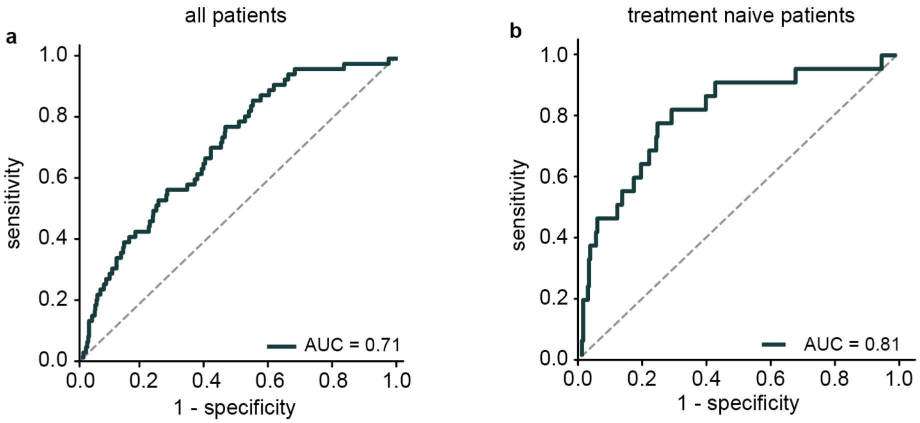


**Figure S2: Time-dependent AUC demonstrates the discriminative performance of ATX for predicting 5-year recurrence risk.** a) Time-dependent ROC curve demonstrating model performance for predicting breast cancer recurrence within a 5-year period across all patients (n = 892). b) Time-dependent ROC curve demonstrating model performance for predicting breast cancer recurrence within a 5-year period across untreated patients (n = 299).

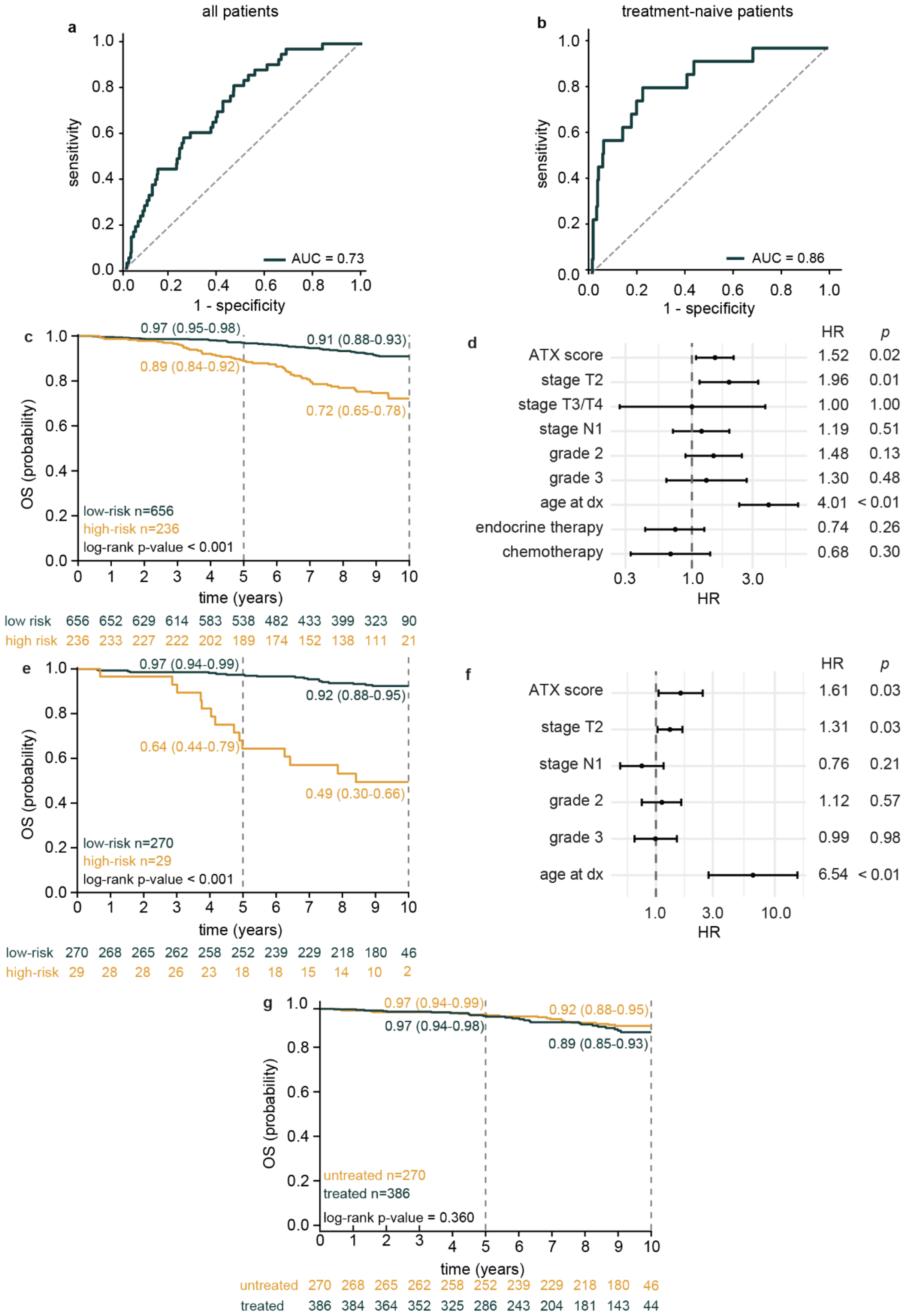
a
all patients
sensitivity
1 - specificity
AUC = 0.73
b
treatment-naive patients
sensitivity
1 - specificity
AUC = 0.86
c
0.97 (0.95-0.98)
0.91 (0.88-0.93)
0.89 (0.84-0.92)
0.72 (0.65-0.78)
OS (probability)
low-risk n=656
high-risk n=236
log-rank p-value < 0.001
time (years)
low risk 656 652 629 614 583 538 482 433 399 323 90
high risk 236 233 227 222 202 189 174 152 138 111 21
d
HR p
ATX score 1.52 0.02
stage T2 1.96 0.01
stage T3/T4 1.00 1.00
stage N1 1.19 0.51
grade 2 1.48 0.13
grade 3 1.30 0.48
age at dx 4.01 < 0.01
endocrine therapy 0.74 0.26
chemotherapy 0.68 0.30
HR
e
0.97 (0.94-0.99)
0.92 (0.88-0.95)
0.64 (0.44-0.79)
0.49 (0.30-0.66)
OS (probability)
low-risk n=270
high-risk n=29
log-rank p-value < 0.001
time (years)
low-risk 270 268 265 262 258 252 239 229 218 180 46
high-risk 29 28 28 26 23 18 18 15 14 10 2
f
HR p
ATX score 1.61 0.03
stage T2 1.31 0.03
stage N1 0.76 0.21
grade 2 1.12 0.57
grade 3 0.99 0.98
age at dx 6.54 < 0.01
HR
g
0.97 (0.94-0.99)
0.92 (0.88-0.95)
0.97 (0.94-0.98)
0.89 (0.85-0.93)
OS (probability)
untreated n=270
treated n=386
log-rank p-value = 0.360
time (years)
untreated 270 268 265 262 258 252 239 229 218 180 46
treated 386 384 364 352 325 286 243 204 181 143 44

**Figure S3: ATX predicts overall survival.** Time-dependent ROC curve demonstrating model performance for predicting overall survival (OS) within a 5-year period across a) all patients (n = 892) and b) untreated patients (n = 299). c) Kaplan-Meier curves illustrating OS stratified by ATX risk category across all patients. d) Multivariate Cox regression model demonstrating ATX is independently associated with OS across all patients. e) Kaplan-Meier curves illustrating OS stratified by ATX risk category across untreated patients. f) Multivariate Cox regression model demonstrating ATX is independently associated with OS across untreated patients. g) Kaplan-Meier plot of OS in ATX-stratified low-risk patients (n = 656).

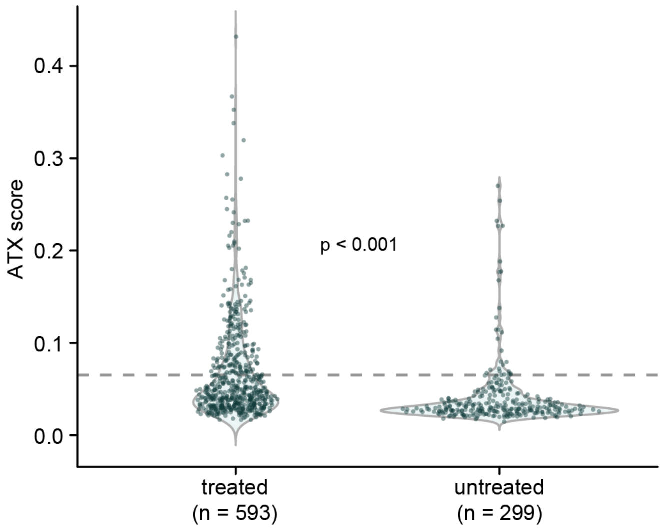


**Figure S4: Distribution of ATX scores by treatment.** ATX scores by treatment group are shown, where each dot represents a single patient. A Wilcoxon rank-sum test found the score distributions differed between the two treatment groups
